# Health Profiles of Factory and Office Workers in Malaysia: A Survey Study

**DOI:** 10.1101/2025.06.12.25329539

**Authors:** Adeola Folayan

## Abstract

**Introduction:** Occupational settings expose workers to varying health risks that may lead to both immediate and long-term health complications. These risks, which include chemical, biological, and physical hazards, can affect individuals differently depending on workplace conditions, exposure levels, and host factors such as immune status. This study aimed to compare the health profiles of workers in three distinct work environments: an electronics factory, an office, and a winery.

**Methodology:** A cross-sectional study was conducted using a self-administered questionnaire that collected data on demographic characteristics, workplace conditions, and self-reported health symptoms. A total of 98 workers with at least six months of experience at their respective workplaces participated. Ethical approval and written informed consent were obtained prior to data collection. Data were analysed using Microsoft Excel 2003.

**Results:** The results revealed distinct health profiles across the three occupational settings. Back and neck pain was commonly reported at all sites, affecting more than 60% of respondents. Electronics factory workers experienced a higher prevalence of cough (62.1%), frequent thirst (45.5%), and headaches (50%), likely linked to chemical exposure and poor air quality. Office workers reported the highest rates of respiratory and skin-related symptoms, including sneezing (71%), dry skin (58.8%), and memory difficulties (47%), suggesting potential indoor air quality issues. Winery workers exhibited a notably higher prevalence of shortness of breath (40%) and frequent thirst (60%), possibly due to poor ventilation and physical exertion.

**Conclusion:** This study provides valuable baseline data on occupational health differences across work settings, underscoring the need for targeted interventions, improved ergonomic designs, and further longitudinal research.

## Introduction

The health profile of a worker may vary due to health problems arising from workplace or job-related risk factors, including exposure to chemical, biological, or physical hazards (1–3). Although everyone is exposed to hazards, not all individuals develop illnesses, as various underlying factors influence disease occurrence. One of the most important factors is the host’s condition. The body has a natural defence mechanism against infectious agents, known as the immune system. However, this immune system may be compromised in hypersensitive individuals, children, the elderly, or people with underlying illnesses (4–6). Furthermore, prolonged exposure to hazards in immunocompetent individuals may eventually suppress the immune system (7). Some of these health symptoms may present as immediate or long-term effects.

This study aims to capture and compare the health profiles of workers across three distinct occupational settings: an electronics factory, an office, and a winery. By examining these diverse work environments, the study seeks to identify potential differences in health outcomes that may be influenced by varying workplace exposures and job-related risk factors.

### Methodology

A self-administered questionnaire was developed based on common symptoms observed in the workplace. The questionnaire comprised sections on demographic information, working conditions, and discomfort complaints, including noise, odour, and temperature.

Prior to the commencement of the study, permission was obtained from the management of the respective study sites. Written informed consent was obtained from each respondent before participation. Ethical approval for the study was granted by the International Medical University’s Ethical Committee. A study information sheet and a written consent form were attached to each questionnaire. Respondents were required to complete the consent form before participating in the study.

The sample size was determined using the Raosoft® sample size calculator, with a 10% margin of error, a 95% confidence interval, and a 50% response distribution, resulting in a minimum required sample size of 96 workers. As the total number of workers at the study sites slightly exceeded this minimum, all willing respondents who met the inclusion criteria were invited to participate. The inclusion criterion required participants to have worked at the study site for at least six months. A total of 98 respondents participated in the study: 66 from the electronics factory, 17 from the office, and 15 from the winery

All data were analysed using Microsoft Excel 2003

## Result

Most respondents were male (77%). The largest age group, constituting 30% of the total respondents, fell within the 23 to 27-year age range, and 56% of the respondents were married. In terms of ethnicity, approximately 39% of the respondents were Chinese, 31% were Malays, 8% were Indians, and the remaining 22% belonged to other ethnic groups.

Back and neck pain was commonly reported across all study sites, with more than 60% of respondents at each site experiencing this symptom. Specifically, 60.6% of workers in the electronics factory, 64.7% of office workers, and 60% of winery workers reported back and neck pain. Office workers reported a higher prevalence of respiratory and skin-related symptoms compared to workers at the other sites. Among office workers, 59% experienced stuffy noses, runny noses, and dry skin, 41% reported nasal irritation, and 71% reported sneezing, which was the highest prevalence of sneezing among the three groups. Memory difficulties were reported by 47% of office workers, compared to 18% of electronics factory workers, while none of the winery workers reported this symptom.

Headaches were reported by 50% of respondents in the electronics factory, 35% in the office, and 47% in the winery. Shortness of breath was reported by 8% of electronics factory workers, 12% of office workers, and 40% of winery workers, with the winery group showing a considerably higher prevalence. Cough was reported by 62.1% of electronics factory workers, 52.9% of office workers, and 26.7% of winery workers. Throat irritation was reported by 27.3% of electronics factory workers, 35.3% of office workers, and 26.7% of winery workers. Frequent thirst was also notable, reported by 45.5% of electronics factory workers, 29.4% of office workers, and 60% of winery workers. Dry skin affected 18.2% of electronics factory workers, 58.8% of office workers, and 26.7% of winery workers. Additionally, skin itchiness was reported by 21.2% of electronics factory workers, 35.3% of office workers, and 26.7% of winery workers.

In summary, electronics factory workers exhibited a higher burden of cough, frequent thirst, headaches, and throat-related symptoms. Office workers experienced the highest rates of sneezing, nasal symptoms, skin discomfort, and memory difficulties. Winery workers showed a notably higher prevalence of shortness of breath and frequent thirst compared to the other groups. The detailed distribution of health symptoms among respondents is presented in Table 1.

**Table 1.** Numbers of respondents with symptoms at the three sites.

|  |  | ELECTRONIC<br>FACTORY | OFFICE | WINERY |
| --- | --- | --- | --- | --- |
| 1 | Shortness of breath | 5 | 2 | 6 |
| 2 | Stuffy nose | 13 | 10 | 2 |
| 3 | Nasal bleeding | 12 | 0 | 1 |
| 4 | Runny nose | 17 | 10 | 3 |
| 5 | Nasal irritation | 5 | 7 | 4 |
| 6 | Sinusous congestion | 11 | 5 | 0 |
| 7 | Sneezing | 40 | 12 | 4 |
| 8 | Cough | 41 | 9 | 4 |
| 9 | Wheezing | 16 | 3 | 0 |
| 10 | Throat irritation | 18 | 6 | 4 |
| 11 | Dry throat | 18 | 7 | 4 |
| 12 | Sore throat | 12 | 6 | 3 |
| 13 | Frequent thirst | 30 | 5 | 9 |
| 14 | Skin itchiness | 14 | 6 | 4 |
| 15 | Dry skin | 12 | 10 | 4 |
| 16 | Skin rashes | 9 | 2 | 0 |
| 17 | Ear irritation | 10 | 1 | 0 |
| 18 | Watery eyes | 10 | 3 | 1 |
| 19 | Headache | 33 | 6 | 7 |
| 20 | Nausea/Vomiting | 10 | 2 | 1 |
| 21 | Diarrhoea | 11 | 1 | 0 |
| 22 | Back and neck pain | 40 | 11 | 9 |
|  | Total number of participants | 66 | 17 | 15 |

## Discussion

It may be difficult to directly associate health complications with a specific health hazard in the workplace, as the mere presence of a hazard does not necessarily confirm it as the cause of the observed health effects (4). Furthermore, identifying the exact causative agent can be challenging since multiple hazards may interact synergistically, producing combined health effects that are difficult to isolate (8, 9). However, this study highlights key differences in the health profiles of workers across three distinct occupational settings: an electronics factory, an office, and a winery.

Back and neck pain emerged as one of the most frequently reported symptoms, with more than 60% of respondents at each site experiencing this complaint. This finding is consistent with existing literature that identifies musculoskeletal discomfort as a common occupational health issue, often linked to poor workstation ergonomics, repetitive tasks, or physically demanding work (10). Employers across all sectors should consider improving ergonomic support and providing more comfortable workstations to mitigate these symptoms.

The electronics factory and winery environments were observed to be noisy, which may explain the high prevalence of headaches reported by workers at these sites. Specifically, 50% of respondents in the electronics factory and 54% of winery workers reported headaches. Prolonged exposure to noise is a well-documented occupational hazard that can contribute to not only hearing loss but also physical and psychological stress, which may manifest as headaches and fatigue (1).

The winery was often stuffy, as noted during the building inspection, which may partially account for the relatively high prevalence of shortness of breath (39%) among winery workers. Exposure to poorly ventilated spaces, combined with fermentation vapours and organic dust, can contribute to respiratory discomfort—a finding that aligns with previous reports from similar occupational settings (11)

Office workers, in contrast, reported higher rates of respiratory and skin-related symptoms. Stuffy noses, runny noses, nasal irritation, dry skin, and sneezing were particularly prevalent, suggesting potential issues with indoor air quality (IAQ) such as poor ventilation, accumulation of dust, or exposure to allergens. These symptoms are commonly associated with “sick building syndrome (12). Additionally, memory difficulties were notably higher among office workers (47%), which may point to a combination of environmental and psychosocial factors, such as cognitive fatigue, poor air circulation, or even chronic exposure to low-level pollutants that could impact cognitive performance (13)

Electronics factory workers experienced a higher burden of cough, frequent thirst, headaches, and throat irritation. These symptoms may suggest exposure to airborne chemicals, dust particles, or vapours commonly associated with manufacturing environments, supporting previous studies that highlight respiratory health risks in such settings (Burton, 2010). Frequent thirst, in particular, may indicate dehydration or exposure to dry air or chemical irritants that contribute to mucosal dryness. While much emphasis has traditionally been placed on chemical contaminants in factory environments, recent studies are also exploring the role of microbial contaminants in contributing to occupational health risks (14–18). Winery workers demonstrated a significantly higher prevalence of shortness of breath and frequent thirst compared to the other groups, which could reflect a combination of physical exertion, exposure to organic particulates, and possible dehydration due to environmental conditions

It is important to note that the cross-sectional design of this study and the reliance on self-reported symptoms limit the ability to establish direct causation. Nonetheless, this study provides valuable baseline data for future research and can serve as a reference point for the development of occupational space guidelines and targeted occupational health interventions. Further studies incorporating environmental monitoring, clinical assessments, and longitudinal follow-ups would enhance the understanding of specific hazard-health relationships in these occupational settings.

## Conclusion

This study has provided important insights into the varying health profiles of workers across three distinct occupational environments: an electronics factory, an office, and a winery. Despite the limitations of the cross-sectional design and reliance on self-reported symptoms, the findings underscore the significance of workplace conditions in shaping workers’ health outcomes. Musculoskeletal discomfort, particularly back and neck pain, emerged as a common issue across all sites, highlighting the urgent need for ergonomic interventions. Additionally, the prevalence of respiratory and skin-related symptoms among office workers suggests potential indoor air quality concerns, while the electronics factory and winery workers appear to face higher exposure to environmental hazards such as noise, airborne chemicals, and poor ventilation.

The distinct symptom patterns observed in each work setting reinforce the importance of tailored occupational health strategies that address the specific risks associated with different job environments. This study serves as a valuable baseline for future research and occupational health policy development, emphasizing the need for improved workplace designs, proactive health surveillance, and regular environmental assessments. Longitudinal studies and detailed exposure measurements are recommended to establish causative relationships between workplace hazards and health outcomes more accurately, ultimately contributing to safer and healthier occupational environments.

## Data Availability

All data produced in the present study are available upon reasonable request to the authors

## Authors’ contributions

AF undertook the literature review, performed the survey and data analysis, and drafted the first version of this paper

## Availability of data

All data produced in the present study are available upon reasonable request to the authors

